# Evaluation of DISORDER: retrospective image motion correction for volumetric brain MRI in a pediatric setting

**DOI:** 10.1101/2020.09.09.20190777

**Authors:** Katy Vecchiato, Alexia Egloff, Olivia Carney, Ata Siddiqui, Emer Hughes, Louise Dillon, Kathleen Colford, Elaine Green, Rui Pedro A.G. Texeria, Anthony N. Price, Giulio Ferrazzi, Joseph V. Hajnal, David W Carmichael, Lucilio Cordero-Grande, Jonathan O’Muircheartaigh

## Abstract

**Background and Purpose:** Head motion causes image degradation in brain MRI examinations, negatively impacting image quality, especially in pediatric populations. Here, we used a retrospective motion correction technique in children and assessed image quality improvement for 3D MRI acquisitions.

**Material and Methods:** We prospectively acquired brain MRI at 3T using 3D sequences, T1-weighted MPRAGE, T2-weighted Turbo Spin Echo and FLAIR, in 32 unsedated children, including 7 with epilepsy (age range 2–18 years). We implemented a novel motion correction technique: Distributed and Incoherent Sample Orders for Reconstruction Deblurring using Encoding Redundancy (DISORDER). For each subject and modality, we obtained 3 reconstructions: as acquired (Aq), after DISORDER motion correction (Di), and Di with additional outlier rejection (DiOut).

We analyzed 288 images quantitatively, measuring 2 objective no-reference image quality metrics: Gradient Entropy (GE) and MPRAGE White Matter Homogeneity (WM-H). As a qualitative metric, we presented blinded and randomized images to 2 expert neuroradiologists who scored them for clinical readability.

**Results:** Both image quality metrics improved after motion correction for all modalities and improvement correlated with the amount of intrascan motion. Neuroradiologists also considered the motion corrected images as of higher quality (Wilcoxon’s z = −3.164 MPRAGE, z = −2.066 TSE, z = −2.645 FLAIR, for all p < 0.05).

**Conclusions:** Retrospective image motion correction with DISORDER increased image quality both from an objective and qualitative perspective. In 75% of sessions, at least one sequence was improved by this approach, indicating the benefit of this technique in unsedated children for both clinical and research environments.

## INTRODUCTION

Head motion is a common cause of image degradation in brain MRI. Motion artifacts negatively impact MR image quality and, therefore, a radiologists’ capacity to read the images, ultimately affecting patients’ clinical care^1^. Motion artifacts are more common in noncompliant patients^2^ but even in compliant adults, intra-scan movement is reported in at least 10% of cases^3^. For children who require high-resolution MR scans, obtaining optimal image quality can be challenging, owing to the requirement to stay still over long durations needed for acquisition^4^. Sedation can be an option, but carries higher risks, costs and preparation and recovery time^5^.

In conditions such as intractable focal epilepsy, identification of an epileptogenic lesion is clinically important to guide surgical treatment. However, these lesions can be visually subtle, particularly in children where subtle cortical dysplasias are more common^6^. Dedicated epilepsy MRI protocols use high-resolution 3D sequences to allow better cortical definition and free reformatting of orientation, but again can involve acquisition times in the order of minutes, so data collection becomes more sensitive to motion^7^.

For children in particular, multiple strategies are available for minimizing motion during MR examinations. Collaboration with play specialists using mock scanners and training or simply projecting a cartoon or movie are good approaches to reduce anxiety^8,9^. These tools are not always an option in clinical radiology and, even with these strategies, motion can still be an issue^10^. Different scanning approaches to correct for this intra-scan motion have been proposed. Broadly, prospective methods track head motion in real-time and modify the acquisition directions accordingly^11^. These approaches are applicable to a wide range of sequences but requires optical systems with external tracking markers, sometimes uncomfortable or impractical, and extra setup can ultimately result in longer examinations. These techniques may also not be robust to continuous motion^11,12,13^. Retrospective techniques have also been proposed, in some cases relying on imaging navigators which are not compatible with all standard clinical sequences or contrasts^12^

Here, we use a more general retrospective motion correction technique: Distributed and Incoherent Sample Orders for Reconstruction Deblurring using Encoding Redundancy (DISORDER). In this method, k-space samples are reordered to enable retrospective motion correction during image reconstruction^14^. Our hypothesis is that DISORDER improves clinical MRI quality and readability. To assess its use for clinical sequences, we acquired a dedicated epilepsy MRI protocol in 32 children across a wide age range. We used both objective image quality metrics and expert neuroradiologist ratings to evaluate the outcome after motion correction.

## MATERIAL AND METHODS

### STUDY POPULATION

We recruited families for a prospective study of pediatric epilepsy (ethics ref 18/LO/1766). Informed consent was obtained from all participants or their parents, as appropriate. From June to November 2019 we recruited 32 participants: 25 healthy controls and 7 children with focal epilepsy, aged 2–18 years (median 11), 16 females (50%) (Table 1). Exclusion criteria were ages under 6 months or over 18 years, major neurological conditions unrelated to epilepsy, and contra-indications for 3T MRI.

**Table 1.** Descriptive demographics of the study population.

| Table 1. Descriptive demographics of the study population. |  |  |  |
| --- | --- | --- | --- |
| Characteristics | Healthy Controls | Patients with Epilepsy | Total |
| Number | 25 | 7 | 32 |
| Age at Scan |  |  |  |
| Mean + SD | 11.32 ± 4.8 | 11.6 ± 3.7 | 11.4 ± 4.5 |
| 1-5 years old | 3 | 0 | 3 |
| 6-10 years old | 9 | 3 | 12 |
| 11-15 years old | 7 | 3 | 10 |
| 16-18 years old | 6 | 1 | 7 |
| Sex |  |  |  |
| Male | 13 | 3 | 16 |
| **Female** | 12 | 4 | 16 |
| *Note. - SD=Standard deviation* |  |  |  |

### IMAGE ACQUISITION

Children were scanned without sedation on a 3T Philips Achieva- TX using a 32-channel head coil. They were asked to stay still during scanning while watching a favorite movie. The protocol was: T1-weighted MPRAGE (repetition time (TR) = 7.7ms, echo time (TE) = 3.6ms, flip angle (FA) = 8°, inversion time (TI) = 900ms, echo train length (ETL) = 154 and acquisition time (TA) = 286s); T2-weighted TSE (TR = 2500ms, TE = 344ms, ETL = 133, TA = 342s); and T2-weighted FLAIR (TR = 5000ms, TE = 422ms, TI = 1800ms, ETL = 182, TA = 510s). Parallel imaging acceleration (SENSE) of 1.4 was used along both phase encoding directions. Field of view was 240×188×240mm and images were 1 mm isotropic. The combined acquisition time was approximately 22 minutes.

All scans were acquired using the DISORDER scheme (Figure 1). A shot of k-space is defined as a portion of k-space phase encoding data in the k_2_k_3_ plane, acquired within a single acquisition block. In Figure 1, each shot is represented by a different color. As demonstrated in ^14^, DISORDER aims to improve motion tolerance by guaranteeing that the acquisition of every shot contains a series of samples distributed incoherently throughout kspace. This is achieved with a modified phase encoding (PE) sampling order. We adopt the “random-checkered” approach illustrated in Figure 1. Data are acquired in the inferior-superior k_1_, anterior-posterior k_2_ and left- right k_3_ orientations, this way rotations on the sagittal plane (k_1_k_2_) are sampled faster within each shot, improving robustness to intra-shot motion. In our protocol the number of shots is 120 (duration 1200ms each) for MPRAGE, 135 (658ms) for TSE, and 100 (859ms) for FLAIR.

**Figure 1:**
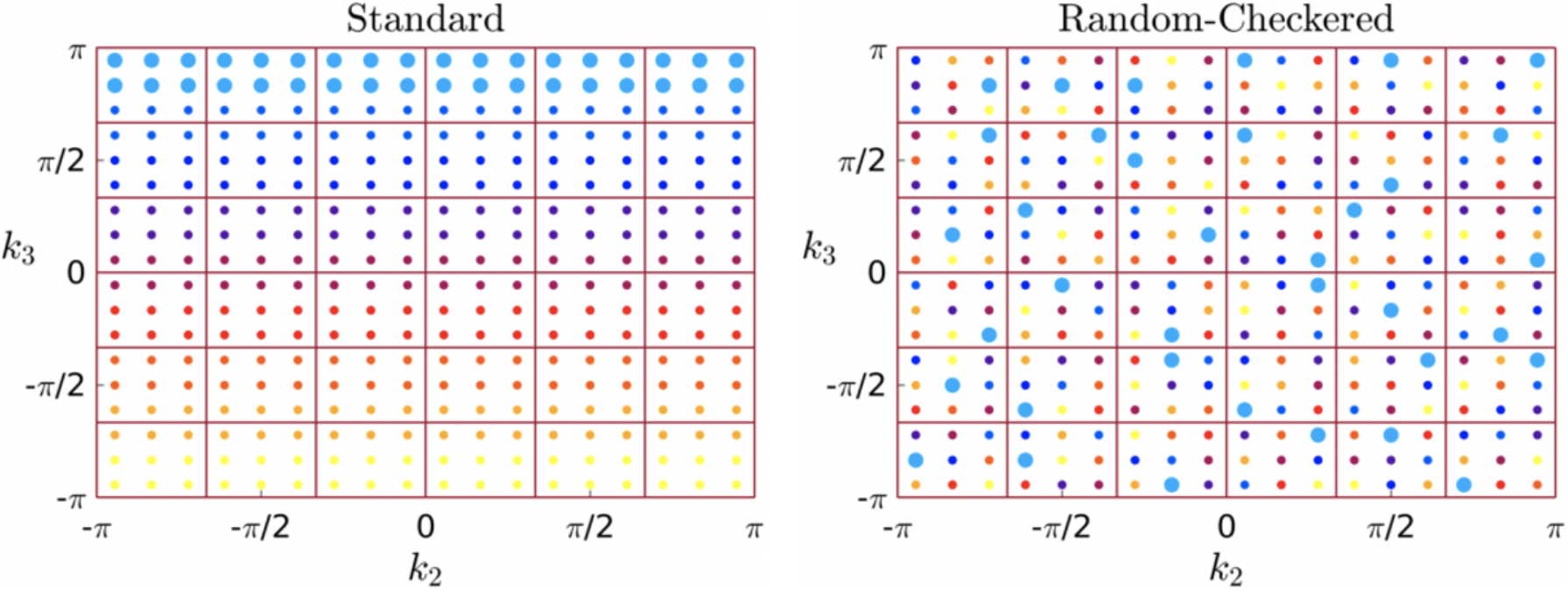
The image shows the different k-space data acquisitions: on the left side the standard acquisition that sequentially acquires adjacent lines in the grid, with an example shot given as bigger dots in blue. The image on the right represents DISORDERs “random checkered” acquisition, in which every shot acquires distributed information in k-space with a certain degree of randomness.

### MOTION CORRECTION

Motion and reconstruction were estimated jointly using a parallel k-space model in the presence of rigid motion^14^. Starting from a standard reconstruction assuming no motion, a first approximation of the motion parameters for each shot is obtained by maximizing the likelihood of the k-space measures for current reconstructed volume. Then, a new volume is reconstructed with current motion parameters and the method alternates between motion estimation and reconstruction until convergence. An in-depth description of the reconstruction algorithm has been described previously^15,14^.

The time for reconstruction varied from 5 to 40 minutes, depending on degree of intra-scan motion. In the case of high intra-shot motion, the DISORDER framework can further improve the image quality by dismissing outlier shots. Therefore, each subject had their images reconstructed in three different ways: as acquired without motion correction (**Aq**), with DISORDER reconstruction (**Di**) and with DISORDER reconstruction including outlier shot rejection (**DiOut**).

Motion estimates from DISORDER correction can also provide a measure of intra-scan motion. To summarize and quantify this intra-scan motion, we averaged the temporal standard deviations of the three rotation parameters (in degrees) for every scan.

### IMAGE QUALITY ASSESSMENT

For all quality assessments there were 288 images available (32 subjects, 3 imaging modalities and 3 reconstructions). To objectively compare image quality, we used two metrics that do not rely on reference datasets: gradient entropy (GE) and white matter signal homogeneity (WMH; MPRAGE only).

The entropy of an image is a measure of sharpness that characterizes its texture based on intensity^15^. GE has been previously used to characterize image definition, smaller when areas of uniform signal intensity are separated by sharp edges^16^. We calculated the normalized GE for Aq, Di and DiOut respectively. Decreased GE indicates that image information is concentrated at the edges, a measure of sharpness. This metric has a high correspondence with visual assessment of clinical MRI^17^.

WMH of the T1- weighted images were obtained using an automatic segmentation in Freesurfer^18^(v6). After calculating a WM mask, the mean and standard deviation (SD) of the signal intensity were computed within the mask. The WMH was then measured as the mean scaled by SD, with higher WMH associated with higher image quality.

Image quality was further visually inspected by two pediatric neuroradiologists with over 9 years’ experience each (A.E and O.C.). Radiology scoring was explicitly for assessment of focal epilepsy which needs very high contrast between grey and white matter. They scored the images with a four-point Likert scale: 1 unreadable (not suited for clinical use), 2 poor quality (main structures identifiable but heavily blurred or artefacts covering more than 50% of the image), 3 good quality (good gray-white matter differentiation, little blurring or minor artifacts), 4 excellent quality (no motion artifacts, good contrast and perfectly defined grey-white matter boundaries). Sagittal, coronal and axial views of all images were presented to the two radiologists on the same screen and room environment in a randomized and blinded fashion. Each rater looked at all 288 cases in several sessions. The image viewer was rview (https://biomedia.doc.ic.ac.uk/software/irtk/). In all reported comparisons (quantitative metrics and quality ranks), Wilcoxon signed-rank tests were used.

## RESULTS

### QUANTITATIVE METRICS

As expected, older children tended to move less than younger ones. This was a consistent relationship, statistically significant for MPRAGE and FLAIR (Spearman’s rho: −0.416, −0.363, both p < 0.05), but not for TSE scans (Spearman’s rho: −0.229, p = 0.21) (Fig 2).

**Figure 2:**
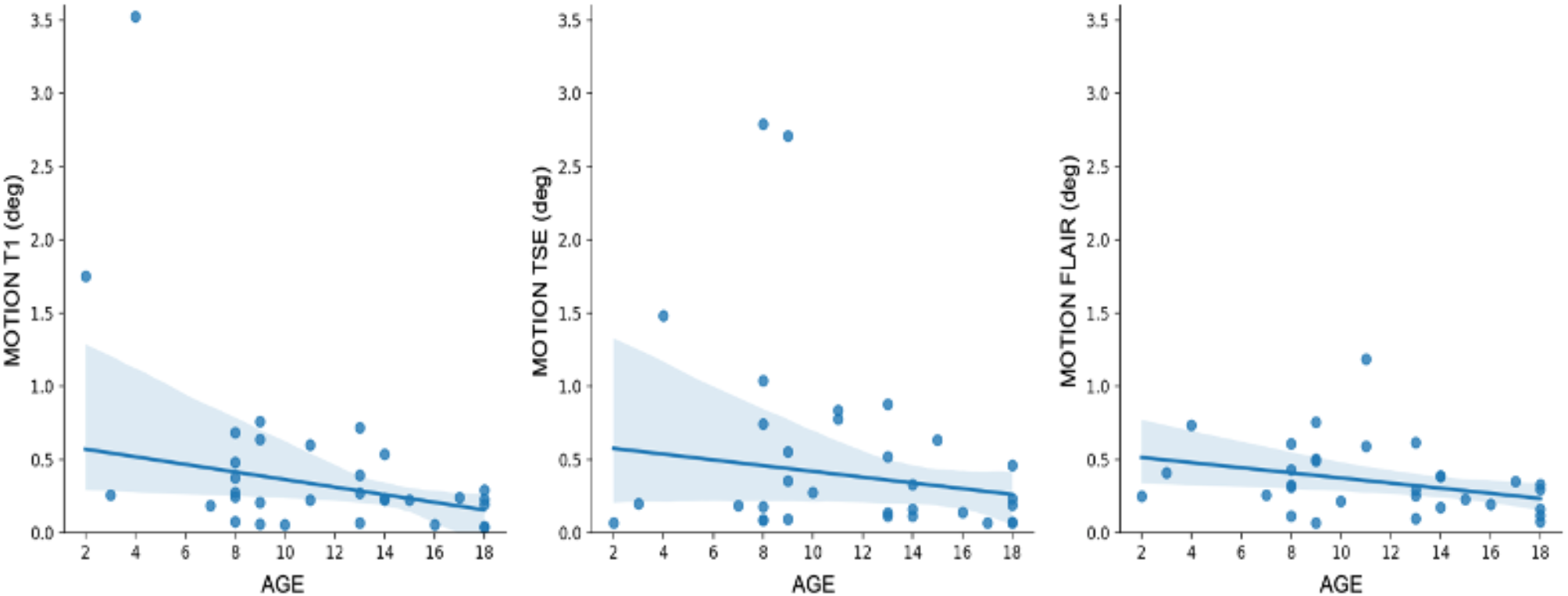
Estimated motion in relation to age. Older children tended to move less than younger ones. This was statistically significant for MPRAGE and FLAIR, but not for TSE.

GE was reduced after motion correction across all modalities: the Wilcoxon’s rank test performed on mean scores before and after motion correction showed a statistically significant difference (for Di z = −4.861 MPRAGE, z = −4.769 TSE, z = −4.884 FLAIR; for DiOut z = −4.937 MPRAGE, z = −4.937 TSE, z = −4.938 FLAIR, for all p < 0.05) (Fig 3).

**Figure 3:**
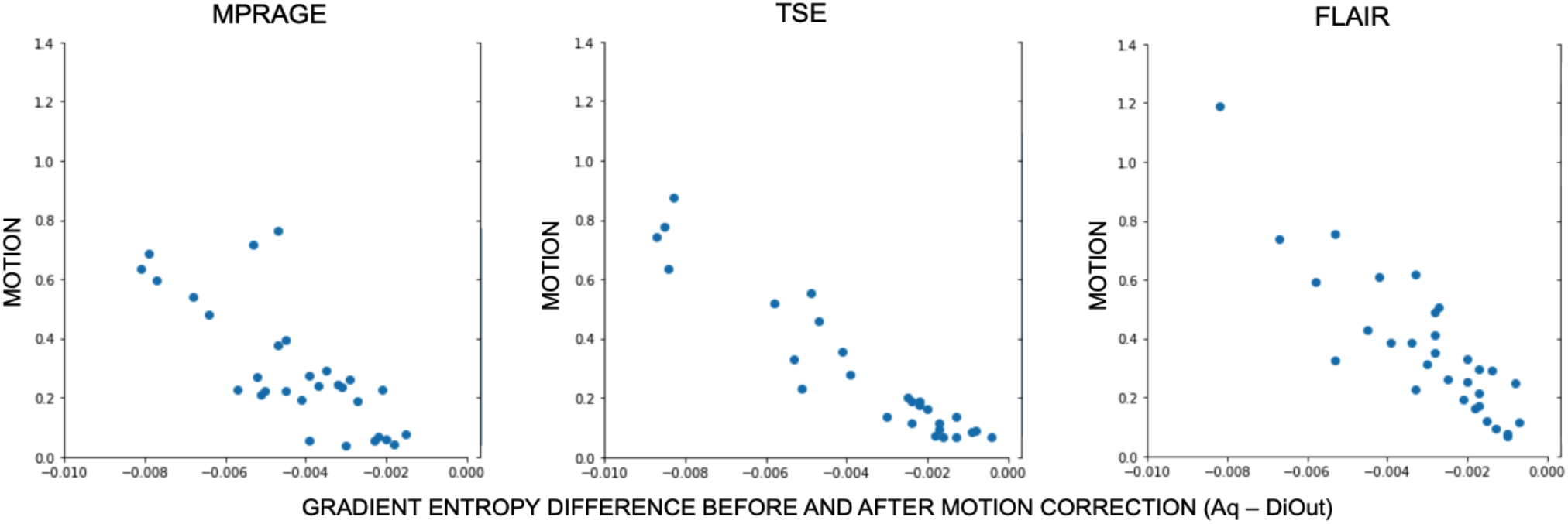
The reduction in gradient entropy by motion correction (DiOut compared to Aq) relative to the estimated amount of motion for every participant in all 3 modalities: from the left MPRAGE, TSE and FLAIR.

There was a linear association between GE decrease after motion correction and degree of intrascan motion. We calculated the difference in GE before and after motion corrected data and estimated a linear regression against motion for each modality. The coefficient of determination (R^2^) for MPRAGE images against motion was R^2^ = 0.24 in Aq-Di, R^2^ = 0.48 in Aq-DiOut; for T2 weighted images is R^2^ = 0.63 in Aq-Di, R^2^ = 0.69 Aq-DiOut; for FLAIR images is R^2^ = 0.44 in Aq-Di, R^2^ = 0.51 in Aq-DiOut, for all p < 0.05. The reduction of GE was larger after outlier rejection (Fig 4).

**Figure 4:**
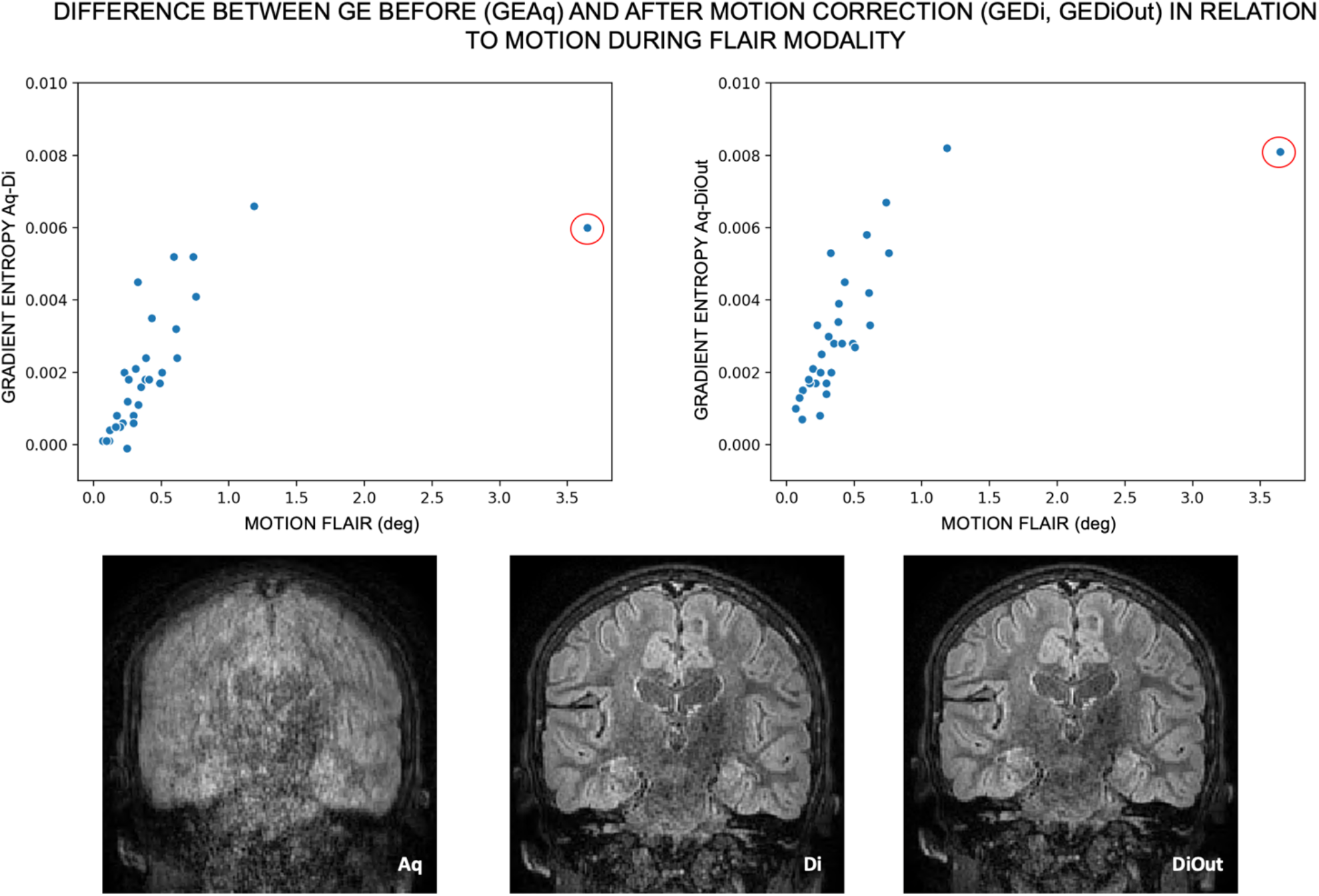
The top row illustrates the difference in gradient entropy before and after motion correction (Di on the left, DiOut on the right) in relation to intra-scan motion for the FLAIR images. The highlighted outlier datapoint (red circle) is shown on the bottom row. The example images show the reconstruction outcome in the subject with the highest intra-scan motion. In this case, GE decreased after motion correction (more in the DiOut image), which visually relates to observers’ score that improved from unreadable (1) in the Aq image, to good and excellent (3/4) in Di and DiOut respectively.

There was an increase of WMH after motion correction on the MPRAGE images (R^2^ = 0.16 for Aq- Di and R^2^ = 0.15 for Aq- DiOut, both p < 0.05). One case was excluded from this analysis due to large motion during the MPRAGE acquisition and a resulting failure of the Freesurfer pipeline for the uncorrected reconstruction. The full brain segmentation was possible on the motion corrected version of the images and WMH was measured (Fig. 5, A).

**Figure 5:**
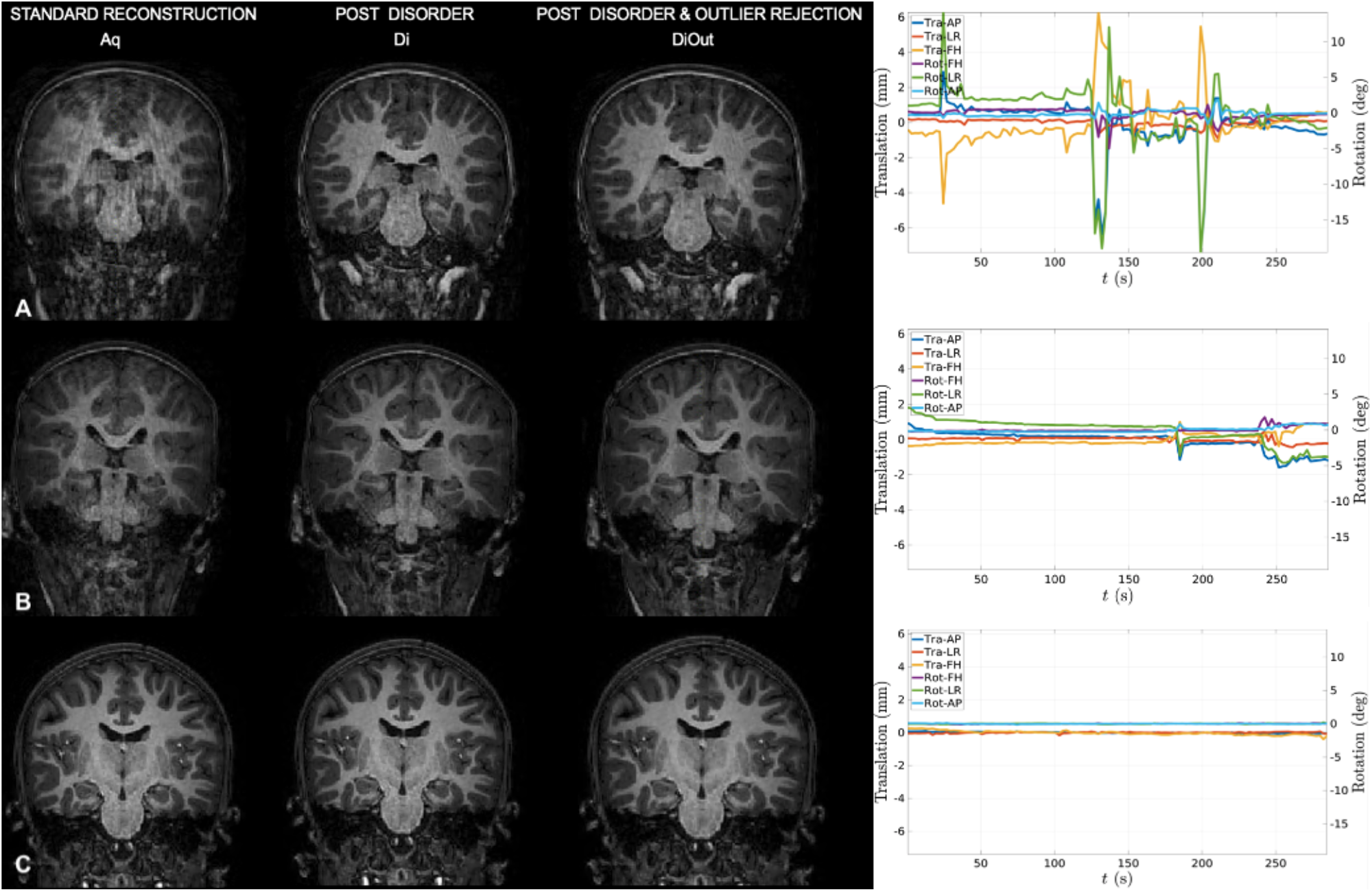
Three individual cases of MPRAGE images before and after motion correction and the corresponding motion trace displaying translations (Tra) and rotations (Rot) in 3 directions: anteroposterior (AP), left-right (LR), foot-head (FH). Respectively: A) high motion,B) moderate motion, C) little/no motion.

### QUALITATIVE METRICS

Expert visual inspection showed that image quality generally improved after motion correction (see Figure 6). There was agreement between observers in raw images scores according to Cohen’s kappa coefficient for inter-rater reliability: k > 0.3 for FLAIR and MPRAGE images, k > 0.6 for TSE images, for all p < 0.05. The intraclass correlation (ICC) of the score change was used as another measure of interrater consistency on rating improvement. The ICC coefficient for absolute agreement in the change in scores after motion correction of the images was > 0.8 for TSE (Di and DiOut) while for FLAIR it was 0.64 for Di and 0.56 for DiOut images, all p < 0.05. The rating increase was less consistent between observers for MPRAGE images: 0.52 (p < 0.05) for Di and 0.37 (p = 0.09) for DiOut.

Wilcoxon signed-rank tests were conducted to compare the expert scores before and after image correction: the improvement of scores was statistically significant both for Di and DiOut and for both radiologists (for observer 1 z = −3.164 MPRAGE, z = −2.066 TSE, z = −2.645 FLAIR, for observer 2 z = −3.162 MPRAGE, z = −2.714 TSE, z = −3.419 FLAIR, all p < 0.05).

The Wilcoxon signed-rank test performed on mean scores between the two types of motion correction (Di and DiOut) did not show any statistically significant difference, except for higher scores for FLAIR DiOut compared to Di (z = −1.97, p = 0.049) for one observer only.

For observer 2, the motion corrected images (Di and DiOut) were all scored equally or higher in comparison to the Aq ones. Observer 1 gave lower ratings for motion corrected reconstructions (Di) compared to acquired in 6/32 cases for FLAIR, 7/32 cases for MPRAGE, 4/32 cases for TSE (by a maximum of 1 point). However, for most scans, the scores increased: 23/32 for FLAIR, 22/32 cases for MPRAGE, 12/32 cases for TSE. For both raters, the score tended to remain good and equal in scans with little or no motion.

As expected, the Spearman’s rho showed a negative correlation between the amount of motion and the resulting score of acquired images in all modalities and for both observers (observer 1: ρ = −0.558 for MPRAGE, ρ = −0.496 for T2 both p < 0.05, ρ = −0.216 for FLAIR, p = 0.24: observer 2: ρ = −0.619 for MPRAGE, ρ = −0.641 for T2, ρ = −0.544 for FLAIR, all p < 0.05).

**Figure 6:**
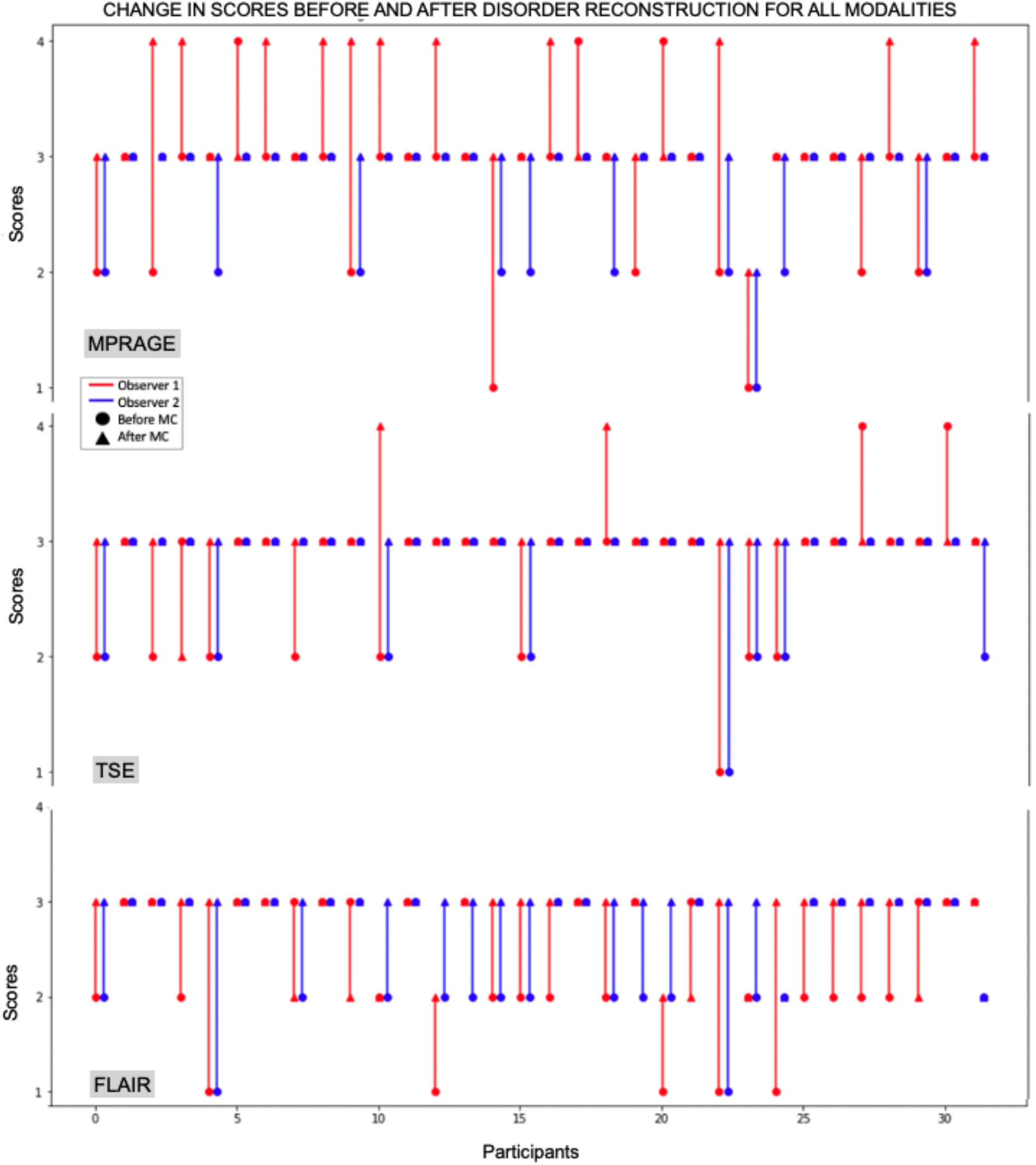
Scores before and after motion correction (DiOut) for all modalities. Blue and red correspond to observer 1 and 2 scores respectively. The dot indicates the score before motion correction, while the triangle indicates the score after correction. Motion correction generally improved the image quality from a radiological perspective.

## DISCUSSION

Successful neuroimaging in childhood is important for both clinical evaluation and research in brain development and disease. However, obtaining high quality data in children is obviously challenging in the highly motion sensitive MRI context^19^. In this work, we demonstrated the benefits of retrospective motion correction on a non-sedated pediatric cohort undertaking brain MR imaging on a 3T scanner using the DISORDER framework. We applied this method to a dedicated high-resolution epilepsy protocol across a wide age range (from toddlers to older adolescents) and showed that DISORDER motion correction increases image quality both quantitatively and qualitatively.

The two neuroradiologist raters broadly agreed on the improved diagnostic value of motion corrected images and, on average, ratings were higher after DISORDER. In 24/32 (75%) participants, at least one modality was improved by DISORDER in a clinically significant way – from being considered unreadable or of poor quality to good or excellent quality. Images acquired in the presence of no or little motion maintained their high quality after motion correction.

In a small number of cases, DISORDER reconstructions were rated lower, though all in the context of very low motion. However, an advantage of this retrospective method is that images both before and after motion correction are always available for radiological evaluation. Observed differences between raters are in line with previous studies^20^ where differences in subjective radiological judgement are reported.

As expected from practical experience and previous studies^21,22^, younger children tended to move more than older ones, which was observed statistically significant for MPRAGE and FLAIR, though not for TSE. None of the participants exhibited very high motion during this acquisition. This was the third sequence acquired in our protocol, and it may be that participants at that time point were simply more been settled or comfortable (engaging with the video they were watching or, in a few cases, spontaneously falling asleep).

The proposed motion compensation method is particularly flexible for use, and it is applicable to any volumetrically encoded sequence. It does require modification of the scanner software to meet the requirements of data acquisition ordering, but it does not involve any additional hardware, significant manipulation of imaging parameters, or additional operator training. The image reconstruction is operated with a vendor-independent off- line open-source code (https://github.com/mriphysics/DISORDER/releases/tag/1.1.0), so that the technique is not restricted to a specific vendor and has been tested on scanners from several manufacturers.

To comply with the requirements of enough SNR for high image resolution (1 mm) and strong motion tolerance, data are acquired with moderate or low acceleration factors (SENSE factor 1.4×1.4). However, the DISORDER encoding does not increase the sequence acquisition time per se. In practice, it may reduce the need for repeat scans and the time overhead would compare favorably with times required for sedation. The method can also provide motion correction for additional 3D sequences where motion correction can be crucial (e.g. relaxometry^23^).

The approach provides clinically useful improvements. In this study, motion correction is applied to a dedicated epilepsy protocol where clinicians are aiming to identify sometime subtle abnormalities such as focal changes in cortical thickness, subcortical signal abnormalities or blurring of the grey-white matter junction. These assessments require time for thorough evaluation. In presence of motion, DISORDER can be a helpful tool as these image features can be enhanced as shown in figure 7.

**Figure 7:**
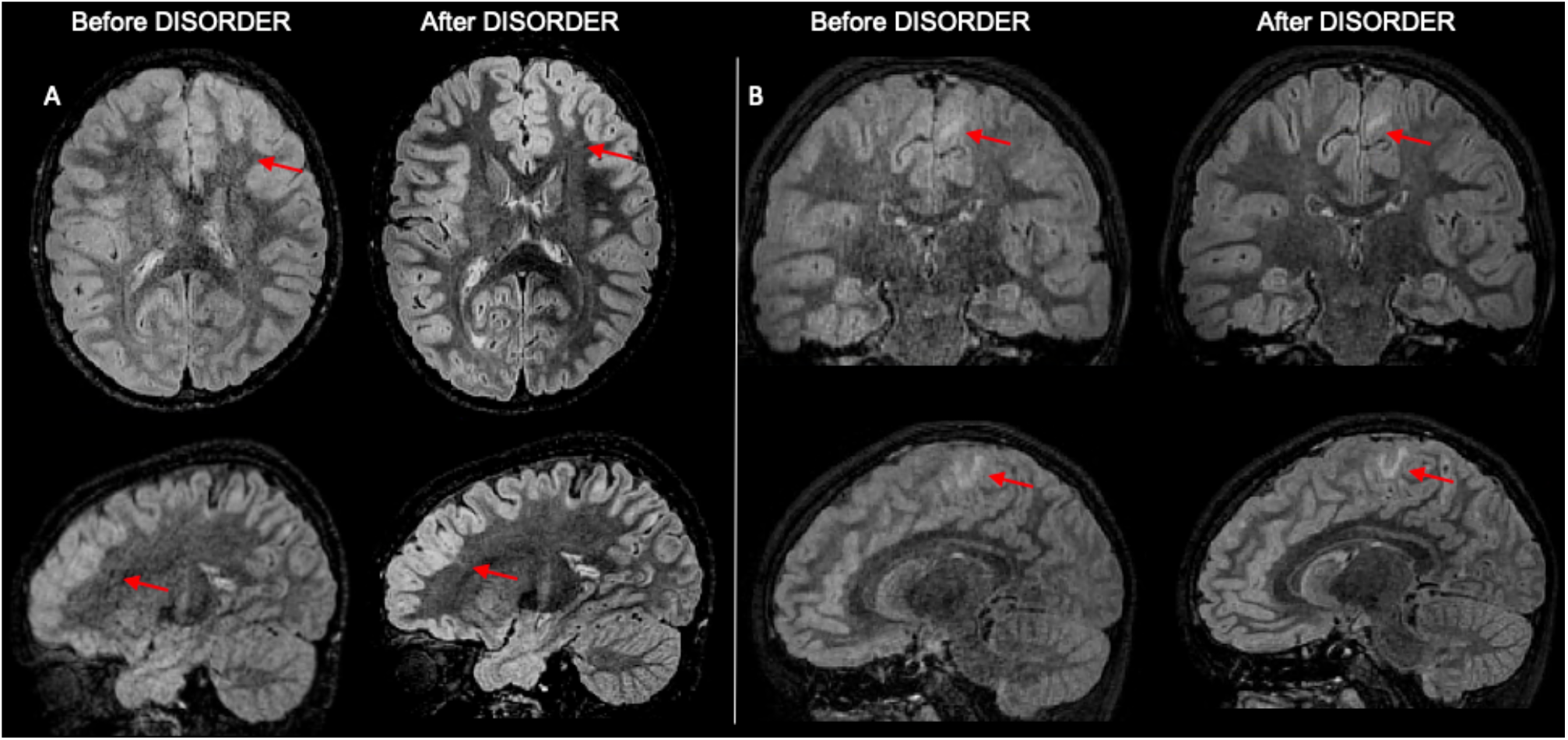
FLAIR images with and without motion correction for two cases. In A the red arrow points toward a tiny area of linear signal abnormality below the left superior frontal sulcus. In B the red arrow highlights an area of focal cortical dysplasia in the left mesial frontoparietal region. These abnormalities are clearly better appreciated after applying DISORDER.

More broadly, DISORDER could be helpful in a clinical setting to improve identification of other types of lesions not necessarily related to epilepsy: notably smaller injuries such as punctate bleeds (where small motion may blur out the injury) or more obvious such as a brain mass (where the extent and edge of pathological tissue can be difficult to discern on blurry images). This method would be beneficial not only for children, but also for patients with high anesthesiologic risk, situations on which time constraints the possibility to repeat scans, and also for adult patients with intellectual disabilities.

Some limitations are noted. First, the method may require longer scan times to perform well in cases of very quick, large range or continuous motion and this isn’t assessed here. Second, DISORDER sampling *increases* motion sensitivity, facilitating its subsequent correction, so some enhancement of artefact levels compared to standard acquisition schemes is likely on the uncorrected images. Two further clinical considerations are also not yet addressed, the impact of reconstruction delay (up to an hour) and a quantification of maximum tolerable degradation before an acquisition needs to be repeated (how bad can the image be before it needs to be repeated). However, given the almost global improvement in data quality for motion corrupted data, the reconstruction delay is probably not a large concern and addressable with future software implementations. Certainly, in this case, no DISORDER reconstructed sequences were considered radiologically unreadable.

This framework for motion-tolerant structural 3D brain images improves clinical MRI image quality both quantitatively and qualitatively. This might have substantial safety and economic implications for healthcare, reducing the clinical indication for sedation and repeat scans in children and adults.

## Data Availability

Code used in this analysis and derived data that support the conclusions of this study are available on request.

## Abbreviations

DISORDER: Distributed and Incoherent Sample Orders for Reconstruction Deblurring using Encoding Redundancy
MPRAGE: Magnetization Prepared Rapid Acquisition Gradient Echo
FLAIR: Fluid Attenuated Inversion Recovery
TSE: Turbo Spin-Echo
GE: Gradient Entropy
WM-H: White Matter Homogeneity

## Bibliography

1. Andre JB, Bresnahan BW, Mossa-basha M, et al. Toward Quantifying the Prevalence, Severity, and Cost Associated With Patient Motion During Clinical MR Examinations. J Am Coll Radiol. 2015;12(7):689–695. doi: 10.1016/j.jacr.2015.03.007

2 Cox AD, Virues-Ortega J, Julio F, Martin TL. Establishing motion control in children with autism and intellectual disability: Applications for anatomical and functional MRI. J Appl Behav Anal. 2017;50 (1):8–26. doi: 10.1002/jaba.351

3 Gedamu EL, Gedamu A. Subject movement during multislice interleaved MR acquisitions: Prevalence and potential effect on MRI-derived brain pathology measurements and multicenter clinical trials of therapeutics for multiple sclerosis. J Magn Reson Imaging. 2012;36(2):332–343. doi: 10.1002/jmri.23666

4 Byars AW, Holland SK, Strawsburg RH, Schmithorst VJ, Dunn RS, Ball WS. Practical Aspects of Conducting Large-Scale fMRI Studies in Children. J Child Neurol. 2002;17(12):885–890.

5 Ho ML, Campeau NG, Ngo TD, Udayasankar UK, Welker KM. Pediatric brain MRI part 1: basic techniques. Pediatr Radiol. 2017;47(5):534–543. doi: 10.1007/s00247-016-3776-7

6 Blumcke I, Spreafico R, Haaker G, et al. Histopathological findings in brain tissue obtained during epilepsy surgery. NEngl J Med. 2017;377(17):1648–1656. doi: 10.1056/NEJMoa1703784

7 Wellmer J, Quesada CM, Rothe L, Elger CE, Bien CG, Urbach H. Proposal for a magnetic resonance imaging protocol for the detection of epileptogenic lesions at early outpatient stages. Epilepsia. 2013;54(11):1977–1987. doi: 10.1111/epi.12375

8 Theiba C, Frayne A, Walton M, et al. Factors associated with successful MRI scanning in unsedated young children. Front Pediatr. 2018;6(May):6–8. doi: 10.3389/fped.2018.00146

9 Dean DC, Dirks H, O’Muircheartaigh J, et al. Pediatric neuroimaging using magnetic resonance imaging during non-sedated sleep. Pediatr Radiol. 2014;44(1):64–72. doi: 10.1007/s00247-013-2752-8

10 Centeno M, Tierney TM, Perani S, et al.Optimising EEG-fMRI for localisation of focal epilepsy in children. PLoS One. 2016;11(2):1–16. doi:10.1371/journal.pone.0149048

11 Maclaren J, Herbst M, Speck O, Zaitsev M. Prospective motion correction in brain imaging: A review. Magn Reson Med. 2013;69(3):621–636. doi: 10.1002/mrm.24314

12 Kecskemeti S, Samsonov A, Velikina J, et al. Robust Motion Correction Strategy for Structural MRI in Unsedated Children Demonstrated with Three-dimensional Radial MPnRAGE. Radiology. 2018;289(2):509–516. doi: 10.1148/radiol.2018180180

13 Stucht D, Danishad KA, Schulze P, Godenschweger F, Zaitsev M, Speck O. Highest resolution in vivo human brain MRI using prospective motion correction. PLoS One. 2015;10(7):1–17. doi:10.1371/journal.pone.0133921

14 Cordero-Grande L, Ferrazzi G, Teixeira RPAG, O’Muircheartaigh J, Price AN, Hajnal J V. Motion-corrected MRI with DISORDER: Distributed and incoherent sample orders for reconstruction deblurring using encoding redundancy. Magn Reson Med. 2020;84(2):713–726. doi:10.1002/mrm.28157

15 Cordero-Grande L, Teixeira RPAG, Hughes EJ, Hutter J, Price AN, Hajnal J V. Sensitivity encoding for aligned multishot magnetic resonance reconstruction. IEEE Trans Comput Imaging. 2016;2:266–280. doi: 10.1109/TCI.2016.2557069

16 Atkinson D, Hill DLG, Stoyle PNR, Summers PE, Keevil SF. Automatic correction of motion artifacts in magnetic resonance images using an entropy focus criterion. IEEE Trans Med Imaging. 1997;16(6):903–910. doi: 10.1109/42.650886

17 Thum CH. Measurement of the entropy of an image with application to image focusing. Opt Acta(Lond). 1984;31(2):203–211. doi: 10.1080/713821475

18 McGee KP, Manduca A, Felmlee JP, Riederer SJ, Ehman RL. Image metric-based correction (Autocorrection) of motion effects: Analysis of image metrics. J Magn Reson Imaging. 2000;11(2):174–181. doi: 10.1002/(SICI)1522-2586(200002)11:2174::AID-JMRI15>3.0.co;2-3

19 Fischl B, Van Der Kouwe A, Destrieux C, et al. Automatically Parcellating the Human Cerebral Cortex. Cereb Cortex. 2004;14(1):11–22. doi: 10.1093/cercor/bhg087

20 Edwards AD, Arthurs OJ. Paediatric MRI under sedation: Is it necessary? What is the evidence for the alternatives? Pediatr Radiol. 2011;41(11):1353–1364. doi: 10.1007/s00247-011-2147-7

21 Tschampa HJ, Kallenberg K, Urbach H, et al. MRI in the diagnosis of sporadic Creutzfeldt-jakob disease: A study on inter-observer agreement. Brain. 2005;128(9):2026–2033. doi: 10.1093/brain/awh575

22 Greene DJ, Koller JM, Hampton JM, et al. Behavioral interventions for reducing head motion. Neuroimage. 2019;(314): 234–245. doi: 10.1016/j.neuroimage.2018.01.023

23 Dosenbach NUF, Koller JM, Earl EA, et al. Real-time motion analytics during brain MRI improve data quality and reduce costs. Neuroimage. 2017;161:80–93. doi: 10.1016/j.neuroimage.2017.08.025

24 Kecskemeti S, Alexander AL. Three-dimensional motion-corrected T1 relaxometry with MPnRAGE. Magn Reson Med. 2020;(December 2019):1–12. doi: 10.1002/mrm.28283

